# Genome Sequencing and Comprehensive Rare Variant Analysis of 465 Families with Neurodevelopmental Disorders

**DOI:** 10.1101/2023.03.01.23285719

**Authors:** Alba Sanchis-Juan, Karyn Megy, Jonathan Stephens, Camila Armirola Ricaurte, Eleanor Dewhurst, Kayyi Low, Courtney E French, Detelina Grozeva, Kathleen Stirrups, Marie Erwood, Amy McTague, Christopher J Penkett, Olga Shamardina, Salih Tuna, Louise C. Daugherty, Nicholas Gleadall, Sofia T Duarte, Antonio Hedrera-Fernández, Julie Vogt, Gautam Ambegaonkar, Manali Chitre, Dragana Josifova, Manju A Kurian, Alasdair Parker, Julia Rankin, Evan Reid, Emma Wakeling, Evangeline Wassmer, C Geoffrey Woods, NIHR-BioResource, F Lucy Raymond, Keren J Carss

**Affiliations:** Department of Haematology, University of Cambridge, UK; NIHR BioResource, Cambridge University Hospitals NHS Foundation Trust, UK; Molecular Neurogenetics Unit, Center for Genomic Medicine, Massachusetts General Hospital, Boston, MA 02114, USA; Department of Neurology, Massachusetts General Hospital and Harvard Medical School, Boston, MA 02114, USA; Program in Medical and Population Genetics, Broad Institute of MIT and Harvard, Cambridge, MA 02142, USA; Centre for Genomics Research, Discovery Sciences, BioPharmaceuticals R&D, AstraZeneca, Cambridge, UK; Clinical Medical School, University of Cambridge, UK; Department of Medical Genetics, University of Cambridge, UK; Centre for Trials Research, Cardiff University, Cardiff, UK; Molecular Neurosciences, Zayed Centre for Research into Rare Disease in Children, UCL Great Ormond Street Institute of Child Health, London, UK; Department of Neurology, Great Ormond Street Hospital for Children NHS Foundation Trust, London, UK; Healx Ltd. Cambridge, UK; Hospital Dona Estefânia, Centro Hospitalar de Lisboa Central, Lisbon, Portugal; Pediatric Neurology Department, Hospital Universitario Central de Asturias, Spain; West Midlands Regional Genetics Service, Birmingham Women’s and Children’s Hospital, Birmingham, UK; Child Development Centre, Cambridge University Hospitals NHS Foundation Trust, UK; Guy’s and St Thomas’ Hospital, London, UK; Department of Clinical Genetics, Royal Devon University Healthcare NHS Foundation Trust, Exeter, UK; North West Thames Regional Genetics Service, Harrow, UK; Neurology Department, Birmingham Women and Children’s Hospital, Birmingham, UK

## Abstract

**Purpose:** Despite significant progress in unravelling the genetic causes of neurodevelopmental disorders (NDDs), a substantial proportion of individuals with NDDs remain without a genetic diagnosis following microarray and/or exome sequencing. Here we aimed to assess the power of short-read genome sequencing (GS), complemented with long-read GS, to identify causal variants in NDD participants from the NIHR BioResource project.

**Methods:** Short-read GS was conducted on 692 individuals (489 affected and 203 unaffected relatives) from 465 families. Additionally, long-read GS was performed on five affected individuals who had structural variants (SVs) in technically challenging regions, complex SVs, or required distal variant phasing.

**Results:** Causal variants were identified in 36% affected individuals (177/489) and a further 26% (129/489) had a variant of uncertain significance, after multiple rounds of re-analysis. Among all reported variants, 88% (333/380) were SNVs/indels, and the remainder were structural variants (SVs), non-coding, and mitochondrial variants. Furthermore, long-read GS facilitated resolution of challenging SVs and invalidated variants of difficult interpretation from short-read GS.

**Conclusion:** This study demonstrates the value of short-read GS, complemented with long-reads, to investigate the genetic causes of NDDs. GS provides a comprehensive and unbiased method to identify all types of variants throughout the nuclear and mitochondrial genome in NDD individuals.

## Introduction

Neurodevelopmental disorders (NDDs) encompass a range of conditions that usually present in childhood, including intellectual disability, developmental delay, autism spectrum disorder, epilepsy, and movement disorders amongst others. While individually rare, collectively NDDs affect millions of people worldwide and present huge challenges for families and healthcare systems.^1^

These disorders are phenotypically and genetically heterogeneous, and are often caused by rare, highly penetrant variants. Over the last decade exome sequencing (ES), and increasingly genome sequencing (GS), have been widely adopted for the identification of NDD-associated pathogenic (P) and likely pathogenic (LP) variants (collectively referred as causal variants throughout this manuscript) in more than 900 NDD-associated genes identified to date.^2,3^ For families with affected children, receiving a genetic diagnosis has many benefits. It often marks the end of a long diagnostic odyssey, can affect clinical management, and allows parents to make more informed subsequent reproductive choices.^1,3^ The proportion of affected individuals in whom a causal variant is identified following genetic testing is known as the diagnostic yield, and it varies according to many factors. For example, in a recent meta-analysis the range of diagnostic yield in studies using ES or GS in children with suspected genetic diseases was 24–68%.^4^

Causal variants are most commonly coding single nucleotide variants (SNVs), small insertions and deletions (indels), and large copy number variants (CNVs).^5^ Additional classes of genetic variation that can cause NDDs include small CNVs (below the resolution of chromosomal microarrays), inversions, translocations, complex structural variants (cxSVs), short tandem repeats (STRs), and variants in the mitochondrial (MT) genome.^1^ Some of these classes of variation are still challenging to detect using short-read sequencing technologies, causing an increasing appreciation of the potential role of long-read sequencing.^6^

The NIHR BioResource has conducted a flagship study whereby short-read GS (srGS) was performed on 13,037 individuals to study the genetic basis of rare disorders including NDDs in the UK national healthcare system.^7^ The aims of the NIHR BioResource NDD study were threefold: 1) to identify a comprehensive range of causal variants using srGS, including those that are often neglected by other methods; 2) to use supplementary long-read GS (lrGS) on a subset to help resolve and interpret variants that were unclear from srGS; 3) to contribute towards the identification of novel NDD-associated genes. This study has been notably successful at achieving all three of these aims. We have contributed towards the identification or confirmation of four NDD-associated genes: *KMT2B, CACNA1E, WASF1*, and *GABRA2*, which have been published elsewhere.^8-11^ In this article we focus on the first two aims and describe for the first time in detail the overall structure and results of the NIHR BioResource NDD study.

## Materials and Methods

### Cohort description

The NIHR BioResource NDD study is a subset of the NIHR BioResource Project,^7^ comprising 692 individuals, of whom 489 were affected with a NDD, and 203 were unaffected relatives. These individuals belonged to 465 families. Recruitment of family members into this study varied depending on availability and suspected mode of inheritance. We sequenced 335 singletons (affected proband only), 67 trios (affected proband and both parents), five quads (affected proband, both parents and a sibling) and 58 families with another family structure combination (Table 1). Most individuals had undergone some negative routine genetic testing prior enrolment to this project, resulting in an enrichment for challenging cases. All participants provided written informed consent and the study was approved by the East of England Cambridge South national institutional review board (13/EE/0325).

**Table 1.** Diagnostic yield by family structure. GS identified causal variants in 36% cases, 23% had a reported VUS and 41% remained unresolved. Partial contribution refers to individuals with a causal variant that partially explains the phenotype. VUS=Variant of Uncertain Significance; LP=Likely Pathogenic; P=Pathogenic. *One trio includes an affected parent.

|  |  |  | Reportable variants: 289 individuals (59%) |  |  | No causal variant identified: 200 individuals (41%) |
| --- | --- | --- | --- | --- | --- | --- |
|  |  |  | P/LP 177 individuals (36%) |  | VUS: 112 individuals (23%) |  |
|  | Family structure | Affected individuals (families) | Full contribution: 168 individuals (34%) | Partial contribution: 9 individuals (2%) |  |  |
| Total affected individuals 489 (465 families) | Singleton | 335 (335) | 111 (33%) | 8 (2%) | 78 (23%) | 138 (42%) |
|  | Trio | 68 (67)* | 28 (41%) | 0 (0%) | 12 (18%) | 28 (41) |
|  | Two siblings | 28 (14) | 16 (57%) | 0 (0%) | 4 (14%) | 8 (29%) |
|  | Cousins | 2 (1) | 2 (100%) | 0 (0%) | 0 (0%) | 0 (0%) |
|  | Proband and parent | 39 (39) | 7 (18%) | 0 (0%) | 14 (36%) | 18 (46%) |
|  | Proband and grandparent | 2 (1) | 0 (0%) | 0 (0%) | 0 (0%) | 2 (100%) |
|  | Quad | 9 (5) | 4 (45%) | 1 (11%) | 2 (22%) | 2 (22%) |
|  | Proband, sibling and parent | 6 (3) | 0 (0%) | 0 (0%) | 2 (33%) | 4 (67%) |

### Short-read GS and identification of causal variants

DNA samples from whole blood underwent short-read GS. Alignment to the human genome of reference GRCh37 and variant calling were performed to identify multiple types of variants including SNVs, indels, structural variants (SVs) and STRs (SFigure 1), as described in the supplementary methods and a previous publication.^7^ Mobile element insertions (MEIs), Spinal Muscular Atrophy (SMA) status and Regions Of Homozygosity (ROHs) were also characterized.

Candidate rare variants were restricted to known NDD-associated genes (see next section) and discussed in multidisciplinary team meetings (MDTs), which included research bioinformatics analysts, clinical scientists and clinical geneticists. Additional information on the variant annotation and filtering strategies is in the supplementary methods. Pathogenicity was determined according to the American College of Medical Genetics guidelines (ACMG).^12^ Causal variants (P/LP) were reported to the patient’s referring clinicians, and variants of uncertain significance (VUS) were reported at the discretion of the MDT. Variants in genes of uncertain association with specific phenotypes were considered for research, further analysis and sharing through Gene Matcher.^13^

### Gene list curation and variant reanalysis

A list of NDD-associated genes was assembled from various sources including OMIM (https://omim.org), DDG2P,^14^ PanelApp^15^ and PubMed searches, then curated to ensure they comply with previously described criteria.^14^ The gene list was updated six times throughout the timeline of the project and the last version contained 1,545 genes (STable 1).

Initially, affected individuals were investigated using the gene list available at the time of analysis. Then, reanalysis of all individuals was performed twice (July 2018 and July 2019) using revised QC and filtering thresholds as well as an updated version of the gene list at the time (v.20180117 and v.20180807 respectively). Re-analysis consisted of manual assessment of 1) rare variants in NDD-associated genes that had been added to the gene list since the first analysis, 2) variants reclassified as P/LP in HGMD or ClinVar since the first analysis and 3) loss-of-function (LOF) SNVs/indels or predicted to be damaging (CADD phred > 20) in NDD-associated genes but with quality metrics below the strict filters employed for the initial analysis. Candidate variants identified by the last approach were manually inspected in IGV (v2.5)^16^ and recommended for Sanger sequencing confirmation.

### Trio analysis

In families where both parents were available (66 trios and 5 quads), joint calling using Platypus variant caller^17^ was also run with default parameters. Then, variants from both algorithms (Platypus and Isaac Variant Caller) were merged, and a gene agnostic identification of candidate variants by mode of inheritance was performed using *in house* filtering scripts described elsewhere.^18^ Variants were interpreted and reported in NDD-associated genes as described above.

### Long-read GS

Long-read GS was done with Oxford Nanopore Technologies (ONT), using the GridION platform for one case (three runs) and the PromethION platform for four cases (four runs). Samples were prepared and sequenced as previously described.^19^ Reads were aligned against the GRCh37 human reference genome and sensitive detection of SVs was performed using and ensemble algorithm approach as previously described.^19^ Additional information on the lrGS methods, algorithms and versions can be found in the supplementary material. Identification of candidate SVs was performed at the locus of interest, and manual inspection of the alignments was also performed using IGV.^16^

## Results

### Diagnostic yield in this NDD cohort achieves 36%

Affected individuals presented with a wide range of NDD phenotypes, and the most frequent were intellectual disability (n=199), seizures (n=191), movement disorders (n=78), dystonia (n=68) and ataxia (n=41), with many individuals having more than one phenotype (SFigure 2). Reportable variants were identified in 59% (289/489) of affected individuals: 36% (177/489) had at least one P/LP variant, and a further 23% (112/489) had at least one VUS (STable 2).

The P/LP variant detection rate was affected by a series of factors. First, diagnostic yield was higher for trios (41%, 28/67) and pair of siblings (57%, 16/28) than probands only (35%, 119/335) (Figure 1A). In four families (STable 2) the reported variants were different amongst multiple affected individuals, supporting previous observations that pathogenic shared variants within the same family should not be assumed.^20^

**Figure 1.**
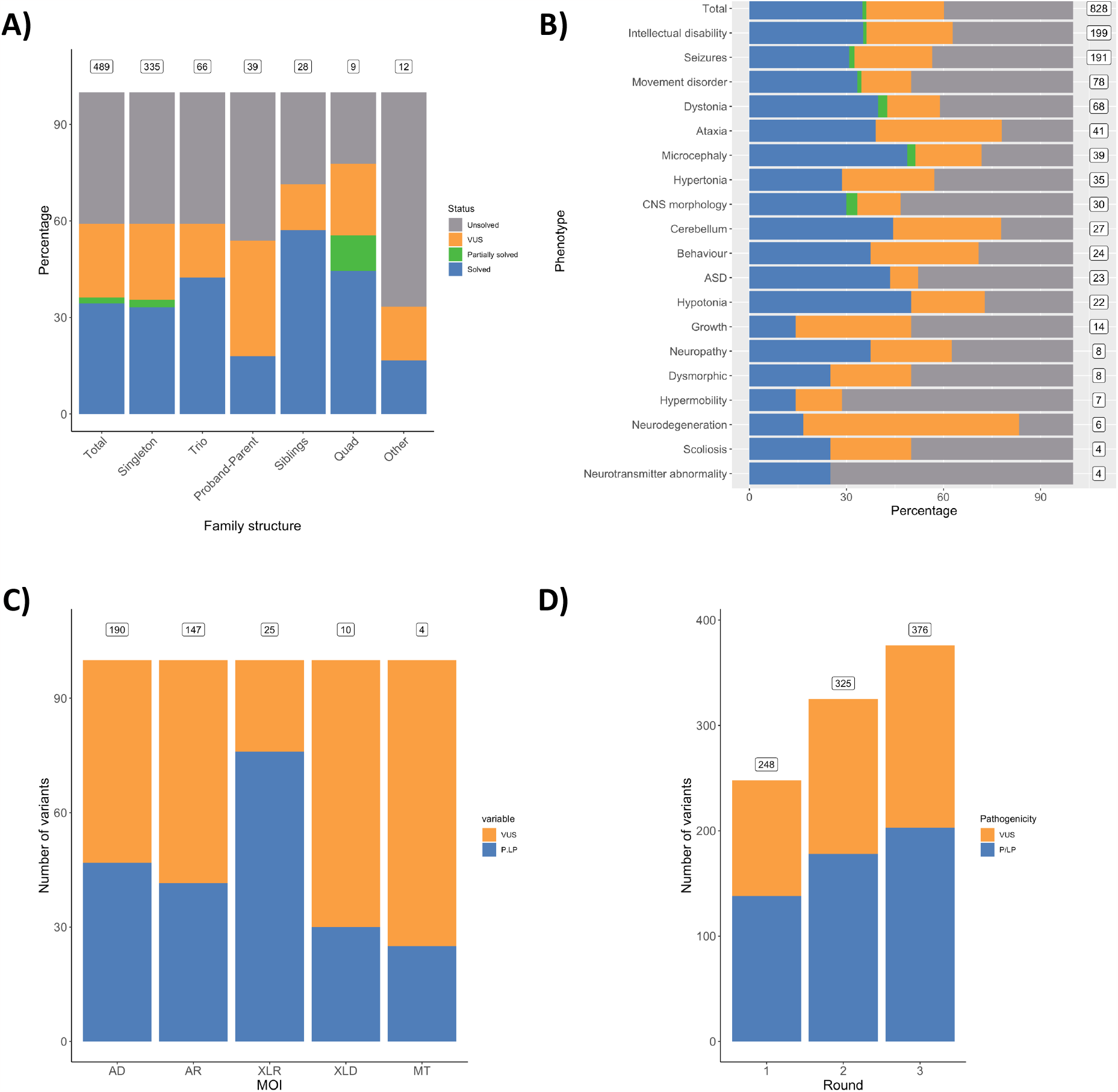
Variant discovery and status rates of affected individuals. **A)** Individual status by family structure and **B)** by phenotype. **C)** Number of variants by mode of inheritance and **D)** by round of analysis. VUS=Variant of Uncertain Significance; ASD=Autism Spectrum Disorder; CNS=Central Nervous System; P=Pathogenic; LP=Likely Pathogenic.

Additionally, diagnostic rate varied depending on genetic ancestry and phenotype (STable 3). While 34% (111/325) of individuals of European ancestry had identified causal variants, only 3% (7/245) of the variants identified in that group were homozygous. The rate was higher in individuals of South-Asian ancestry, where 40% (29/72) of the variants were homozygous and 43% (35/82) of individuals had P/LP variants, which was consistent with previously reported results ^21^.

Furthermore, phenotypes with higher diagnostic rates include hypotonia (50%, 11/22), microcephaly (49%, 19/39), cerebellum abnormalities (44%, 12/27) and autism spectrum disorder (43%, 10/23) while abnormality of growth (14%, 2/14) and hypermobility (14%, 1/7) were lower (Figure 1B). Seventeen individuals with reportable variants had comorbidities and four of these had variants in multiple genes, each partially explaining the phenotype.

### A wide variety of reportable genes and variants are identified in this cohort

The most frequently reported gene across families in the whole cohort was *GNAO1* [MIM: 139311] (*n*=7), followed by *CACNA1A* [MIM: 601011] (*n*=6), *KCNQ2* [MIM: 602235] (*n*=6), *STXBP1* [MIM: 602926] (n=6) and *SCN1A* [MIM: 182389] (*n*=6) (STable 4). In total we reported 380 variants in 276 families, with 21 occurring in more than one family, so overall we identified 358 unique events. The majority of these were SNVs (74%, 279/380), indels (14%, 54/380) and deletions (8%, 31/380) (Table 2). Although duplications, insertions, complex SVs and ROH were found in a lower frequency, in total they accounted for 4% (16/380) of the reported variants. (Figure 1C, STable 5). Although mosaic variants were not systematically called due to the coverage, five likely mosaic variants were identified in this cohort after evaluation of allelic balance and visual inspection of candidate variants in IGV: three were SNVs and two were SVs (STable 2, SFigure 3).

**Table 2.** Candidate variants identified by pathogenicity and type. SNV=Single Nucleotide Variant; SV=Structural Variant; ROH=Region Of Homozygosity; STR=Single Tandem Repeat; SMA=Spinal Muscular Atrophy; VUS=Variant of Uncertain Significance.

| Type | Total | Pathogenic | Likely pathogenic | VUS |
| --- | --- | --- | --- | --- |
| **SNV** | 279 | 48 | 84 | 147 |
| **Indel** | 54 | 23 | 22 | 9 |
| **Deletion** | 31 | 7 | 16 | 8 |
| **Duplication** | 6 | 0 | 0 | 6 |
| **Complex SV** | 4 | 0 | 2 | 2 |
| **Large Insertion** | 3 | 0 | 0 | 3 |
| **Inversion** | 2 | 0 | 0 | 2 |
| **ROH** | 1 | 1 | 0 | 0 |
| **STR expansions** | 0 | 0 | 0 | 0 |
| **SMA** | 0 | 0 | 0 | 0 |
| **Total** | **380** | **79** | **124** | **177** |

The proportion of variants reported as P/LP compared to VUS varied according to variant type. While this proportion was similar for SNVs, 83% of indels (45/54) and 74% of the reported deletions (23/31) were labelled as P/LP. Duplications, large insertions and inversions were all reported as VUS (*n*=11), reflecting the more challenging interpretation of variant effect. One ROH was identified in an individual with Angelman syndrome and deemed to be pathogenic. No STR expansions in known locus or SMA-associated variants were identified in this cohort, which was unsurprising since most of these individuals have previously had a negative routine genetic testing.

### Re-analysis of the data increases diagnostic yield

The first round of variant analysis took place between March 2016 and January 2018. During this time the gene list was under active development, and probands were analyzed using the most recent gene list version available at the time. Reanalysis of the data was performed twice, considering updated variant annotations, quality filtering strategies, and NDD-associated genes. Reanalysis in July 2018 and July 2019 increased the number of reportable variants from 264 to 328 then to 380 respectively (Figure 2D), and it substantially increased affected individuals with reportable variants: from 42% (208/489) to 59% (289/489) after 18 months.

**Figure 2.**
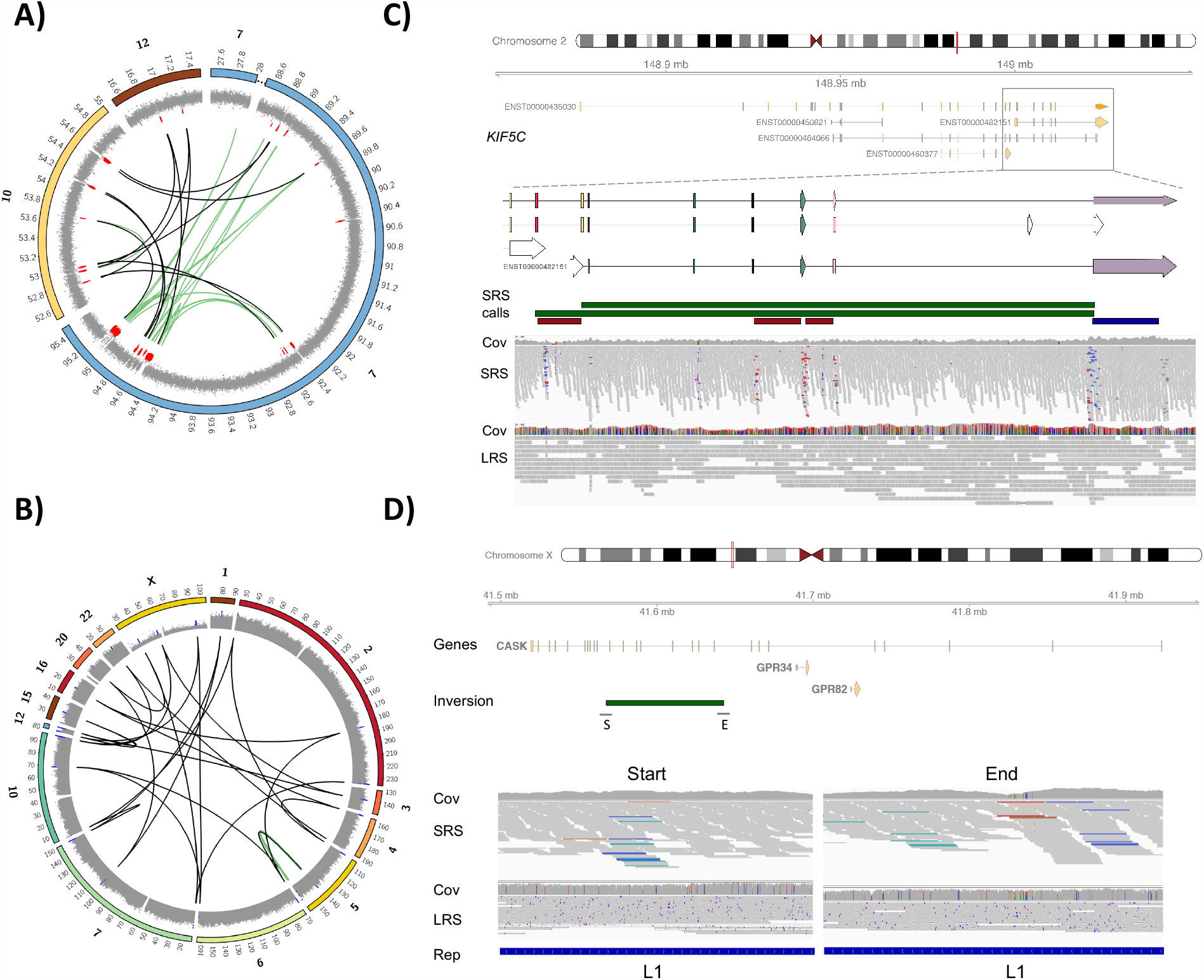
Complex structural variants resolved by lrGS. **A)** Complex rearrangement in Case 6, involving 37 breakpoints between chromosomes 7, 10 and 12. **B)** Complex rearrangement in Case 7, involving 26 duplicated fragments from 14 chromosomes. **C)** Variant phasing performed on Case 10 facilitated resolution of a complex event involving a retroelement of *KIF5C* gene. **D)** Variant phasing performed on Case 8 demonstrated the absence of an inversion called in the short-read sequencing data. Cov=Coverage; SRS=Short-read Sequencing; LRS=Long-read sequencing; Rep=Repeats.

Reanalysis identified new reportable variants due to a variety of reasons: most were in novel NDD-associated genes (69%, 79/115) or were identified due to improvements in the pipeline (28%, 32/115), such as better transcription prioritization, inclusion of MEIs, ROH, or improved *de novo*/SV calling. For example, a variant in *PNPLA6* gene [MIM: 603197] (Chr19:g.7619929 C>T in G008170) was flagged as low quality in the SNV/indel pipeline (minimum overall pass rate of 0.98%), but manual evaluation in IGV suggested it was real; a compound heterozygous variant in *BRAT1* [MIM: 614506] was reported in one case after new publications revealed stronger phenotypic evidence; and one individual had a deep intronic variant in *TSC2* [MIM: 191092] that was identified after it was reported in ClinVar. Additionally, 3.5% (4/115) of variants were in autosomal recessive genes with a previously identified single event, highlighting the importance for analyzing not only novel genes, but also previously known that may harbor missed clinically relevant variants.

### GS detects classes of variants that may be missed by other technologies

Variants that are often challenging to detect by routine diagnostic technologies such as ES and chromosomal microarrays analysis (CMA) include SVs, rare intronic variants, and MT variants. Here we describe findings involving these types of variants in this cohort and we briefly describe ten cases to highlight the value of GS. Additional information for each case and variant is present in the supplementary material and STable 2.

Regarding SVs, we reported a total of 31 deletions, six duplications, two inversions, three large insertions and four cxSV. Importantly, 65% (24/37) of them were smaller than standard CMA resolution (200 Kbp using Affymetrix Chromosome Analysis Suite), underscoring the value of GS to detect SVs cryptic to CMA. Six SVs occur in conjunction with a SNV/indel in a known recessive gene. One example is Case 1, an individual with Early Infantile Epileptic Encephalopathy (EIEE) and a combination of an inversion and a missense variant in *SPATA5* [MIM: 613940], which is associated with an autosomal recessive neurodevelopmental disorder that often includes seizures (SFigure 4). This example underscores the value of GS to investigate inversions, which are often neglected in genetic analyses.

Six intronic variants identified in this cohort were associated with NDDs: five splice region and one deep intronic variant. The latter was in an individual with Tuberous Sclerosis who had endured a long diagnostic odyssey (Case 2). A heterozygous deep intronic variant in *TSC2* [MIM: 191092] was identified in 17% (4/23) of the reads, suggesting mosaicism (SFigure 3b), later confirmed by Sanger sequencing. This variant was observed during reanalysis, after it was published and submitted to ClinVar as associated with disease.^22^

Lastly, four reportable variants were identified in MT genes, three of which were likely pathogenic. Variants were called at different levels of heteroplasmy (from 83-91%) and homoplasmy, which were estimated from coverage analyses. One example (Case 3) is an individual with ataxia, recurrent lactic acidosis and myopathy. This patient had a missense variant in heteroplasmy (91% in blood), in *MT-TL1* gene (SFigure 5). This is one of the most thoroughly studied and best characterized disease-causing MT variants, and is associated, amongst other phenotypes, with MELAS (myopathy, encephalopathy, lactic acidosis, and stroke like episodes),^23^ which was consistent with the patient’s phenotype. The other two LP variants (Case 4 and 5) were in the genes *MT-ATP6*, associated with neurogenic muscle weakness, ataxia and retinitis pigmentosa,^24^ and *MT-ND4*, associated with Leber Hereditary Optic Neuropathy,^25^ respectively (SFigure 6 and 7).

### Long-read sequencing resolves complex SVs in two individuals

Five individuals with ambiguous results from srGS data were further investigated by ONT lrGS (Table 3). A total of seven runs (three in GridION for one sample and four in PromethION for the remainder) produced an average coverage of 14.6 (± 7.5) reads with an average length of 4,243 bp (± 4,054) (SFigure 9A-D). After QC, 62,620 SVs were identified, an average of 26,311 ± 4,532 per individual (SFigure 9E-F), which is consistent to previously reported lrGS studies.^26^

**Table 3.** **Long-read GS** was performed on five cases to resolve cxSVs, variant phasing and to facilitate resolutions of technically challenging regions in five individuals.

| Individual | Phenotype | Finding srGS | Reason inclusion lrGS | Finding lrGS |
| --- | --- | --- | --- | --- |
| **Case 6**<br>**(NGC00375_01)** | Dystonia, myoclonus; delayed gross motor development; learning and intellectual disability | cxSV involving *SGCE* gene | Unable to resolve by srGS, highly complex | cxSV involving 37 breakpoints |
| **Case 7**<br>**(G012664)** | Paroxysmal intermittent limping right leg; bulbar palsy | cxSV involving multiple duplications | Unable to resolve by srGS, highly complex | cxSV involving 26 duplicated fragments |
| **Case 8**<br>**(G013428)** | Severe global developmental delay; hypotonia with chorea like movement disorder; sensorineural hearing impairment; microcephaly; delayed visual maturation with esotropia | Inversion chrX:41426631-41501873 | Unable to resolve by srGS and Sanger sequencing | Variant not supported by lrGS |
| **Case 9**<br>**(G013407)** | Early infantile epileptic encephalopathy | ENSP0000036201 4.3:p.Arg361Pro | Haplotype phasing | Inconclusive |
| **Case 10**<br>**(G000973)** | Early onset dementia; spastic paraplegia; thin corpus callosum | cxSV involving *KIF5C* gene | Unable to resolve by srGS, possible complex retrotransposon | Retrotransposon insertion in chr5:25000434 – not complex, unknown effect |

Two patients carried complex SVs that were resolved by lrGS. Case 6, a male with dystonia, learning difficulties and behavioral problems, had a *de novo* complex SV disrupting *SGCE* [MIM: 604149], which is associated with dystonia. Short-read GS had suggested this was part of a complex SV, but resolution could not be achieved due to homology at the breakpoints. Long-read GS allowed SV characterization and resolved the complex rearrangement that involved 37 breakpoints between chromosomes 7, 10 and 12 (Figure 3A). The variant was reported as LP.

Case 7 is a male with paroxysmal dyskinesia and bulbar palsy, who harbored a complex rearrangement characterized by the presence of duplications across multiple chromosomes, including chromosome X. The variant had been inherited from the unaffected mother and lrGS revealed 26 duplicated DNA fragments of 24Kb median size (sd ± 12Kb) from 14 different chromosomes (Figure 3B). Although no protein coding gene was predicted to be disrupted, we couldn’t rule out the possible regulatory effect of this event, and it was classified as VUS.

### Long-read sequencing phases variants and facilitates resolution of technically challenging regions in three individuals

LrGS was also used to perform variant phasing and to investigate SVs in technically challenging regions. Case 8 (G013428) presented with global developmental delay, hypotonia with movement disorder, sensorineural hearing impairment, microcephaly and delayed visual maturation with esotropia. An inversion involving *CASK* [MIM: 300172] gene was called in the srGS data (Figure 3D), but the variant couldn’t be confirmed by long-range PCR due to low sequence complexity. We therefore sought to validate it using lrGS, and the inversion was not supported by the lrGS data, indicating that the called inversion was a false positive.

Case 9 was a female with EIEE and a heterozygous missense variant in *DNM1* gene [MIM: 616346], which is associated with epileptic encephalopathy. The variant was absent in the unaffected father, and maternal DNA was unavailable (SFigure 8). Given that 80% of *de novo* variants occur in the paternal allele,^27^ we performed lrGS to determine the haplotype of the variant. Unfortunately, the closest informative SNV was 7,048 bp from this position and there were no reads of this length covering the region (average read length 6,723 bp ± 4,695 bp). Therefore, the variant was classified as VUS.

Lastly, Case 10 was a female with early onset dementia, spastic paraplegia and thin corpus callosum. Three deletions and two inversions were called in *KIF5C* [MIM: 604593]. LrGS was used to resolve this event and demonstrated that *KIF5C* had not been disrupted and the calls were from a retroelement insertion of a *KIF5C* transcript highly expressed in human brain (Figure 3C). Although the insertion was not affecting any protein coding gene, it was classified as VUS since reports have shown that retroelements can interfere with gene expression by other mechanisms such as silencing by transcriptional or RNA interference.^28^

## Discussion

In this study we describe in detail the structure and outcomes of the NIHR BioResource NDD project. We employed a comprehensive approach that combined short and long-read GS to identify a broad range of clinically relevant variants associated with NDDs. This strategy identified a high rate of causal variants throughout the nuclear and mitochondrial genomes (36%), including variants often intractable to ES/CMA. Our diagnostic yield is within the expected range reported by similar studies,^3,4,29^ and 3% higher than the 33% reported in a previous NIHR BioResource study due to reanalysis and follow up studies.^7^ It is worth noting that the diagnostic yield for NDDs can vary considerably and is influenced by many factors, such as phenotype and recruitment criteria, sequencing technology, mode of inheritance, family members studied, date of analysis, and genetic ancestry. Understanding these factors can help inform recruitment strategies and study design to improve diagnostic yield.

A notable strength of this study is how comprehensively we surveyed multiple types of variants that could be implicated with NDDs. We not only investigated coding SNVs and indels, but also explored SVs, intronic variants, STR expansions, SMA status and MT variants. However, we did not find any pathogenic STRs or SMA cases, which could be due to several reasons: i) some participants may have undergone STR expansion/SMA testing prior to enrollment, resulting in a reduced likelihood of detecting such variants, ii) these are very rare causes of NDDs, and thus our study may have been underpowered to detect them, and iii) it is possible that these types of variants are identified with lower sensitivity than other classes, or they may be specifically implicated in phenotypes poorly represented within this study.

Interpretation of variants that are not SNVs or indels, such as SVs, can be particularly challenging, despite recent improvements on guidelines for interpretation of CNVs.^30^ Pathogenic intronic and other ‘non-coding’ variants are rare and difficult to identify and interpret, especially without supporting transcriptomic data from an appropriate tissue.^7,31^ Large-scale genome sequencing cohorts currently underway will help improve our understanding of the distribution, features, and function of non-coding variants, facilitating easier identification of those that are pathogenic.^3,7,32,33^ Classes of variants that we were unable to investigate in this study include those in repetitive regions that are intractable to detection by srGS, as well as somatic or mosaic variants that generally require higher coverage sequencing for detection.

Interestingly, we have identified causal variants in several clinically actionable genes. Five individuals have pathogenic variants in *KMT2B* [MIM: 606834]; so may be responsive to treatment with deep brain stimulation.^8,34^ Five other cases have causal variants in *SCN1A* [MIM: 182389]; in these patients treatment with sodium channel blockers can worsen seizures.^35^ These examples demonstrate the clinical importance of genetic diagnoses and the value of this study.

Reanalysis of sequencing data notably increased the diagnostic yield, largely due to causal variants identified in genes newly associated with NDD, as has previously been reported.^29^ This is an important argument for GS or ES over panel sequencing. The decision of whether to reanalyze data for any given cohort must balance this advantage against the resource required, and it will depend partly on number of novel gene-disease associations.

The use of lrGS in human genomics has expanded greatly over recent years, largely due to technological improvements along with development of new algorithms for processing and interpreting the data.^36^ Applications include insights into the biology and consequences of SVs^26,37^ and identification of pathogenic variants in rare diseases that were intractable to other methodologies, usually in individual cases.^6,19,38,39^ Here, we used lrGS to resolve complex SVs that could not be characterized by short-reads in two individuals, and to validate or phase variants in three additional cases. Haplotype phasing in Case 9 was not possible due to read-length limitation, highlighting the importance for ultra-long reads.^38,40^ Overall, our results give several examples of the utility of long-read sequencing. In the future, larger-scale, more systematic lrGS studies of NDDs, facilitated by further improvements to technology, algorithms and pipelines, will yield further insights into the prevalence and biology of previously intractable pathogenic variants.

Our work demonstrates the value of GS to investigate the genetic basis of NDDs and provides insight into the genetic architecture of these disorders. We support the importance of re-analysis and demonstrate that variants cryptic to traditional technologies such as small and cxSVs, non-coding and MT variants can be captured by GS increasing diagnostic yield. Further detailed characterization of genomic variation in large-scale GS studies will be essential for further unveiling the genetic architecture of NDDs in coding and non-coding regions of the human genome.

## Data Availability

Genome data has been deposited at the European Genome-phenome Archive (EGA) under accession number EGAD00001004522.

https://ega-archive.org/datasets/EGAD00001004522

## Acknowledgements

We thank the participants involved in this study and their families.

## Author contributions

Conceptualization: A.S.-J., K.C., F.L.R.; Data curation: A.S.-J., K.M., L.C.D., K.C.; Formal analysis: A.S.-J., K.C.; Funding acquisition: NIHR, F.L.R.; Investigation: A.S.-J., K.M., C.A.R., K.L., C.E.F., D.G., L.C.D., K.C., F.L.R.; Methodology: A.S.-J., K.C., F.L.R.; Project administration: K.S., E.D., M.E.; Resources: A.M.T., S.T.D., A.H.-F., J.V., G.A., M.C., D.J., M.A.K., A.P., J.R., E.R., E.W., E.W., C.G.W.; Software: A.S.-J., C.P., O.S., S.T., N.G.; Supervision: K.C., F.L.R.; Validation: J.S.; Visualization: A.S.-J.; Writing-original draft: A.S.-J., K.C.; Writing-review & editing: A.S.-J., K.C., F.L.R.

## Ethics declaration

All participants provided written informed consent to participate in the study. The study was approved by the East of England Cambridge South national institutional review board (13/EE/0325). The research conforms with the principles of the Declaration of Helsinki. Written informed consent to participate was obtained to publish clinical information.

## Additional information

The online version of this article contains supplementary material.

