## Supplementary material for "Genome Sequencing and Comprehensive Rare Variant Analysis of 465 Families with Neurodevelopmental Disorders"

##### Affiliations

### Index

*Authors* 1

*Supplementary Methods* 3

**Short-read genome sequencing** 3

**Long-read genome sequencing** 3

*Supplementary Figures* 4

*Supplementary Results* 7

Case 1: G013396 7
Case 2: G004131 8
Case 3: G004703 8
Case 4: G013808 9
Case 5: G012198 9
Case 6: NGC00375_01 9
Case 7: G012664 10
Case 8: G013407 10
Case 9: G000973 11
Case 10: G013428 11

*Supplementary references* 13

### Supplementary Methods

#### Short-read genome sequencing

DNA samples from whole blood underwent short-read genome sequencing (GS) using an Illumina HiSeq 2500 or HiSeq X instrument, as previously described in detail (1). Reads were aligned to the human reference genome build GRCh37 using Isaac Aligner (2), and SNVs and indels were called using Isaac Variant Caller (2). The median coverage was 40x (sd ± 3.93) for autosomes and 3,339x (sd ± 1,510) for the MT genome. Quality control (QC) analyses were performed as part of the NIHR BioResource Project and included variant quality checks, coverage analysis and sex and ancestry prediction (1).

Variant annotation and filtering were performed using *in-house* workflows as described elsewhere (1). Briefly: SNVs/indels were annotated using Ensembl's Variant Effect Predictor (VEP) (3) and filtered for population frequency (Minor Allele Frequency (MAF) < 0.01 in ExAC/gnomAD (4), version r2.0.2) and consequence ('Moderate' or 'High' impact). Variants previously reported as pathogenic in HGMD (version PRO 2018.1) (5) or Clinvar (2018-07-29) (6) with a MAF <= 0.1 were also considered, independently of the consequence.

Structural variants (SVs) were identified using a combination of two algorithms: Canvas (v.1.1.0.5) (7), , and Manta (v.0.23.15) (8). cxSVs were identified using Bedtools cluster and categorized as previously described (9). Only rare SVs (estimated sample overlap frequency of < 0.01 in the NIHR BioResource) were investigated. Detailed information regarding data processing and variant filtering have been reported elsewhere (1, 9).

Other types of variation were also studied: STR expansions were called at specific known disease-associated loci using ExpansionHunter (v2.5.5) (10); Spinal Muscular Atrophy (SMA) were identified with SMA caller (v1.0) (11); and mobile element insertions (MEIs) were called using MELT (v2.1.5) (12). All algorithms performed internal variant annotation and were run with default parameters. Additionally, Regions of Homozygosity (ROH) were identified from genomic VCF files using PLINK (v1.9) (13), with optimized parameters for GS data (14). Only regions previously associated with Uniparental Disomy (UPD) disorders were reviewed.

#### Long-read genome sequencing

Long-read GS was done with Oxford Nanopore Technologies (ONT), using the GridION platform for one case (three runs) and the PromethION platform for four cases (four runs). Samples were prepared and sequenced as previously described (15). After basecalling with Guppy (version 4.0.11+f1071ce), reads were aligned against the GRCh37 human reference genome using minimap2 (2.17-r941) (16) with default parameters for nanopore data ('-ax map-ont' parameter). Sensitive SV discovery was done using a combination of three different algorithms: Sniffles (v1.0.11) (17), NanoSV (v1.2.4) (18) and SVIM (v1.2.0) (19), and merged as previously described (15). Identification of candidate SVs was performed at the locus of interest, and manual inspection of the alignments was also performed using IGV (20). Variant phasing for one of the cases was performed with marginPhase (1.0.0, https://github.com/benedictpaten/marginPhase).

### Supplementary Figures

**SFigure 1. Short-read genome sequencing and analysis workflow**. Comprehensive variant identification was performed in all individuals, followed by variant filtering and interpretation according to the ACMG guidelines (21).

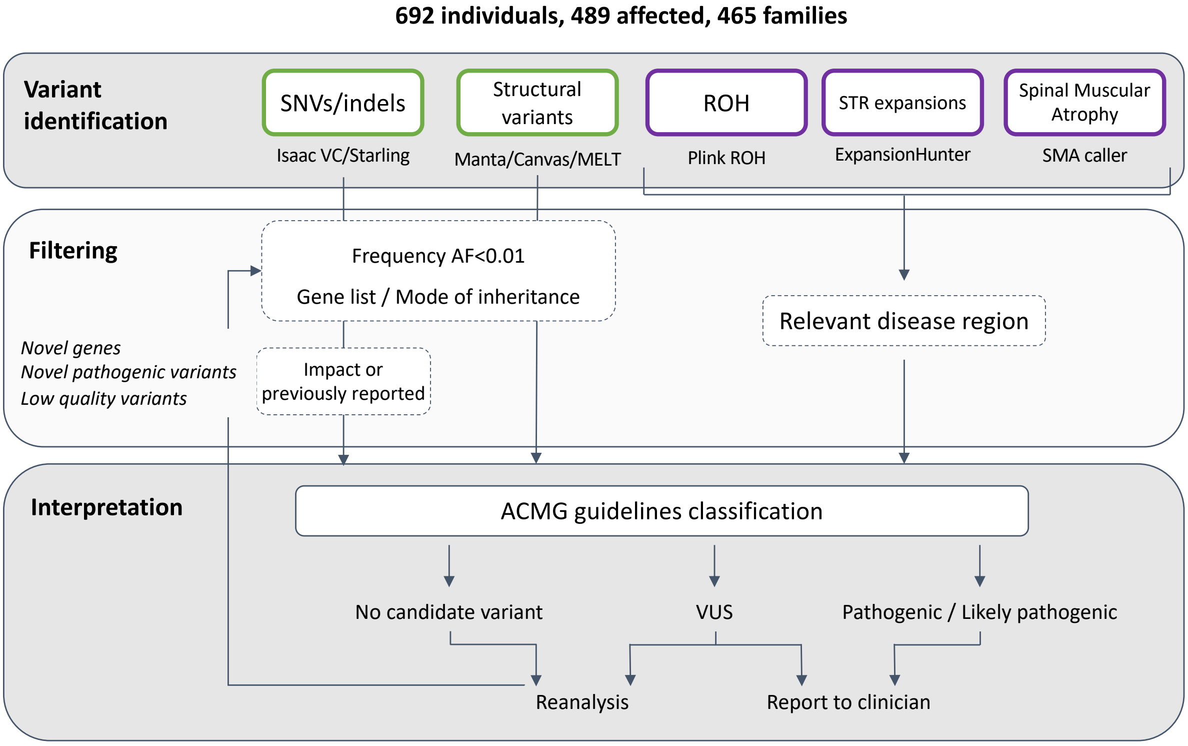


**SFigure 2. Phenotype distribution in the NIHR BioResource NDD study.** The most common phenotypes in this cohort are intellectual disabilities, seizures, movement disorders and other abnormalities of the nervous system. Comorbidities are also present in some individuals.

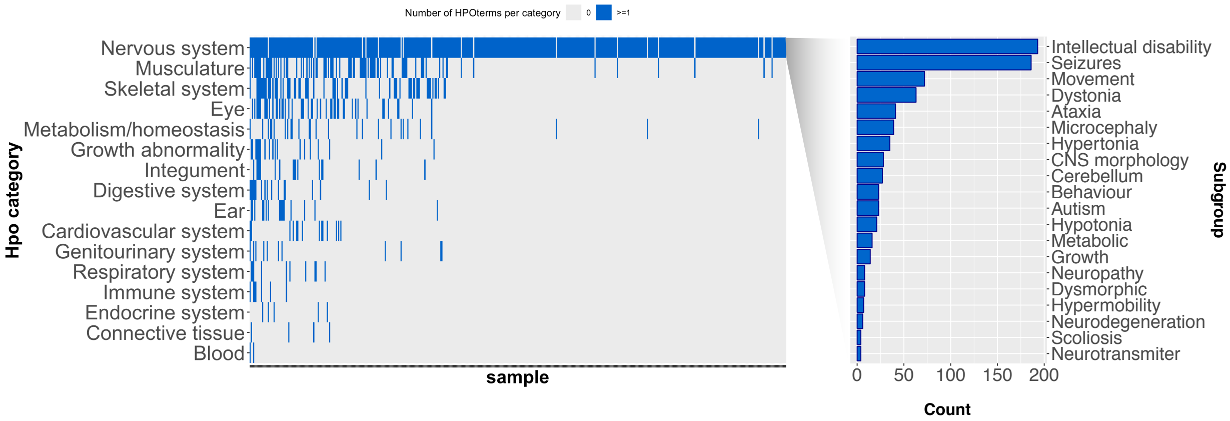

**SFigure 3. Reads supporting likely mosaic variants. a)** Mosaic SNV identified in *SCN2A* [MIM: 182390] in G001312, an individual with intellectual disability (ID), seizures and movement disorder; **b)** Deep intronic mosaic variant in *TSC2* [MIM: 191092] identified in G004131, an individual with Tuberous Sclerosis; **c)** Stop gain SNV in *TRIP12* [MIM: 604506] identified in G004705, an individual with ID and developmental delay; **d)** Mosaic deletion identified in G003412, an individual with dystonia; **e)** Mosaic deletion in a non-pseudoatosomal region of chromosome X in a male individual with cerebellar abnormalities (G011510). The deletion encompassed the last two exons of *KDM6A* [MIM: 300128] and was absent in the mother of this patient. Average coverage within the deleted region was 6x in the proband and 36x in the mother, while for the proband the coverage was 18x and 0x in the pseudoautosomal and non-pseudoautosomal regions of chromosome X respectively.

**a)**

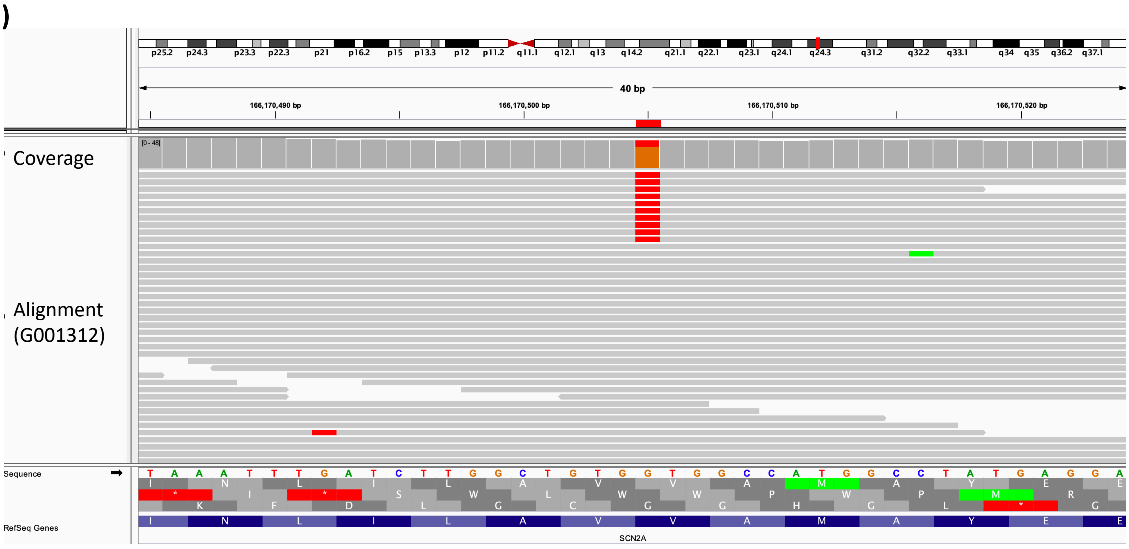


**b)**

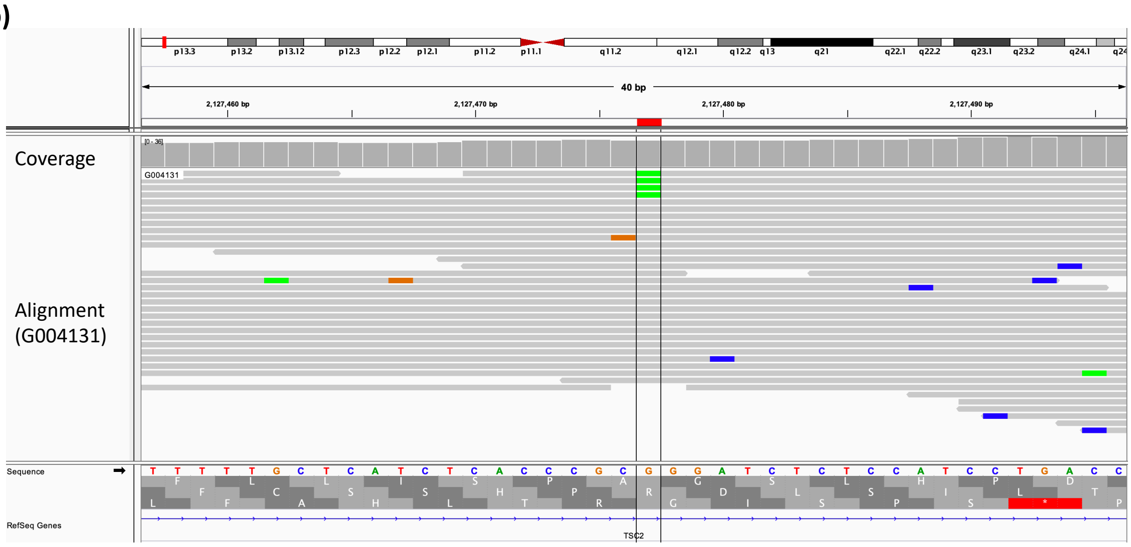

c)

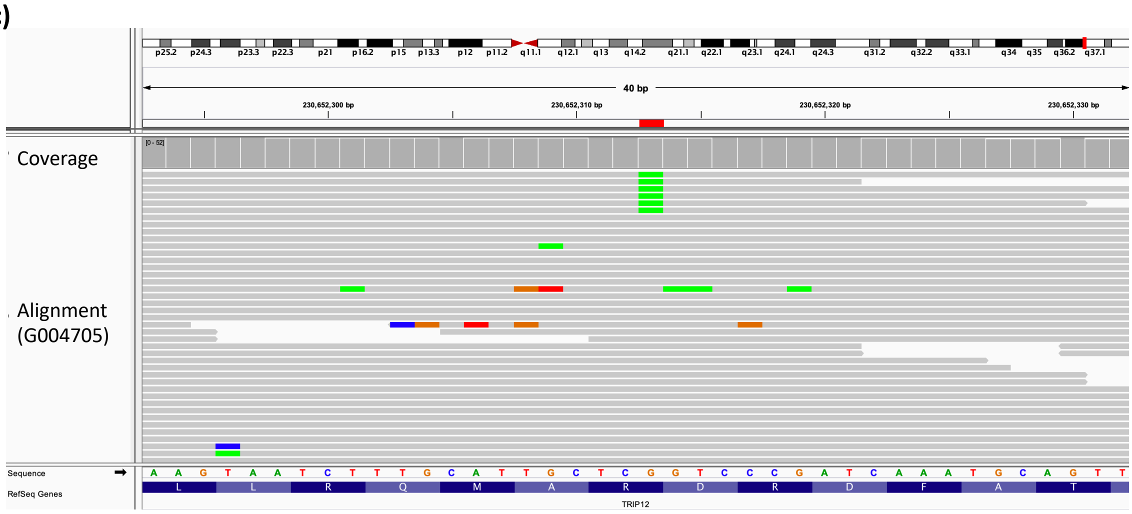
40 bp
230,652,300 bp
230,652,310 bp
230,652,320 bp
230,652,330 bp
Coverage
Alignment
(G004705)
Sequence
RefSeq Genes
TRIP12

d)

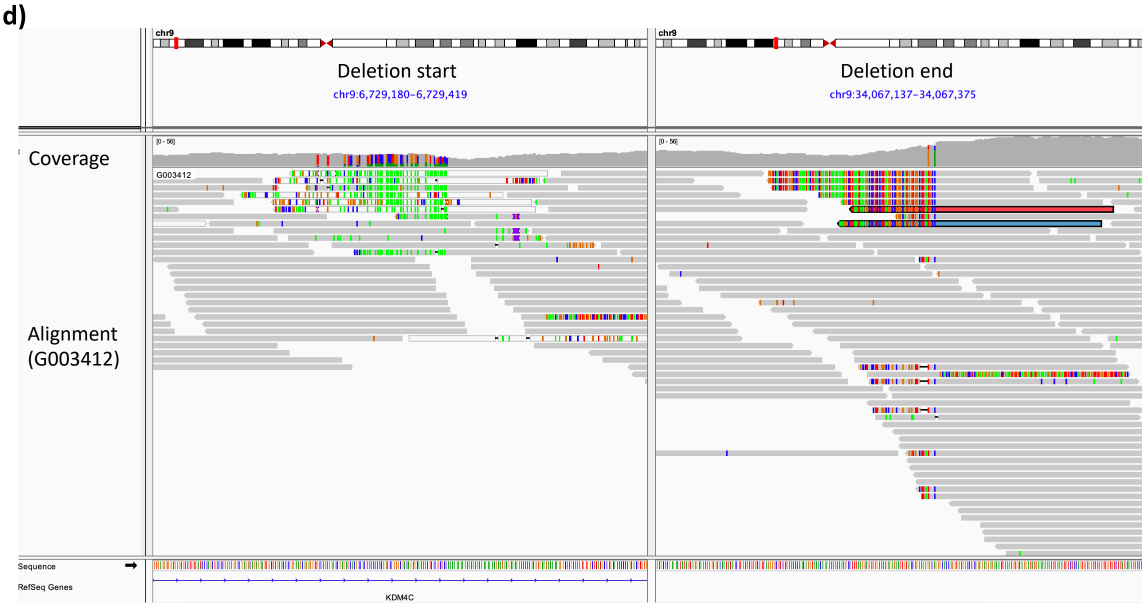
chr9
chr9
Deletion start
chr9:6,729,180-6,729,419
Deletion end
chr9:34,067,137-34,067,375
Coverage
G003412
Alignment
(G003412)
Sequence
RefSeq Genes
KDM4C

e)

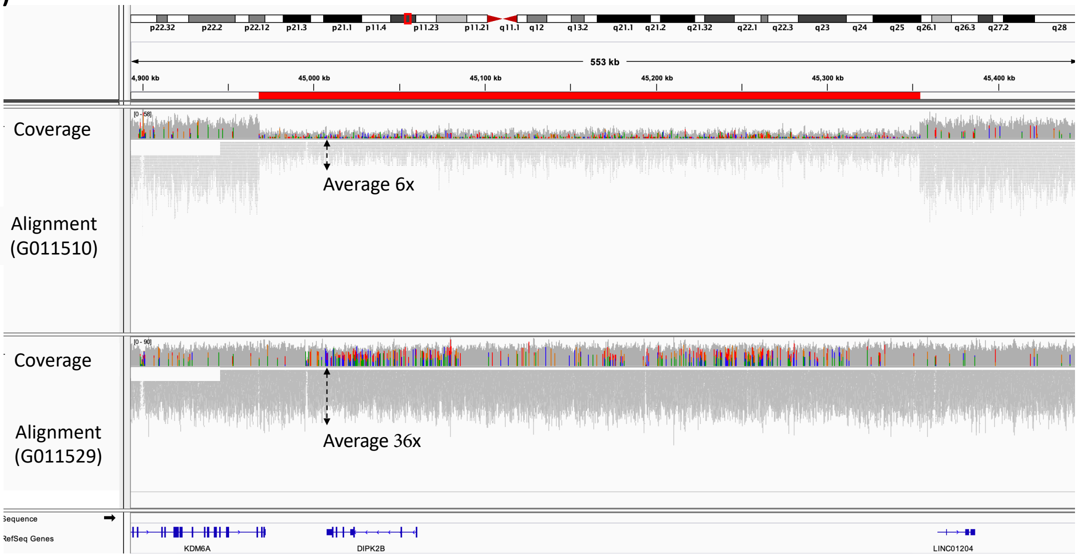
553 kb
44,900 kb
45,000 kb
45,100 kb
45,200 kb
45,300 kb
45,400 kb
Coverage
Alignment
(G011510)
Average 6x
Coverage
Alignment
(G011529)
Average 36x
Sequence
RefSeq Genes
KDM6A
DIPK2B
LINC01204

#### Supplementary Results

##### Case 1: G013396

Individual with Early Infantile Epileptic Encephalopathy (EIEE) and a combination of an inversion and a missense variant in *SPATA5* [MIM: 613940], which is associated with an autosomal recessive neurodevelopmental disorder that often includes seizures. The inversion (Chr4:g123328096-124088220) was inherited from the unaffected mother and the missense variant was unique to this individual. The variants were deemed to be VUS because 1) paternal DNA was unavailable for segregation analysis to confirm compound heterozygosity and 2) although the missense variant (Chr4:g.123949428T>G, ENST00000274008.4:c.1957T>G, ENSP00000274008.3:p.Phe653Val) is absent in gnomAD and predicted to be damaging by multiple in-silico algorithms (CADD phred = 31, SIFT = 0, Polyphen = 1), it had not been reported prior to this publication. Nonetheless, this example underscores the value of genome sequencing (GS) to investigate the role that usually neglected variants such as inversions may play in the development of neurodevelopmental disorders (NDDs), alone or in conjunction with other variants.

**SFigure 4. a)** Inversion involving *SPATA5* in Case 1 with EIEE (G013396), observed to be present in the mother (G013423). **b)** Missense variant in *SPATA5* is present in the proband and absent in the mother.

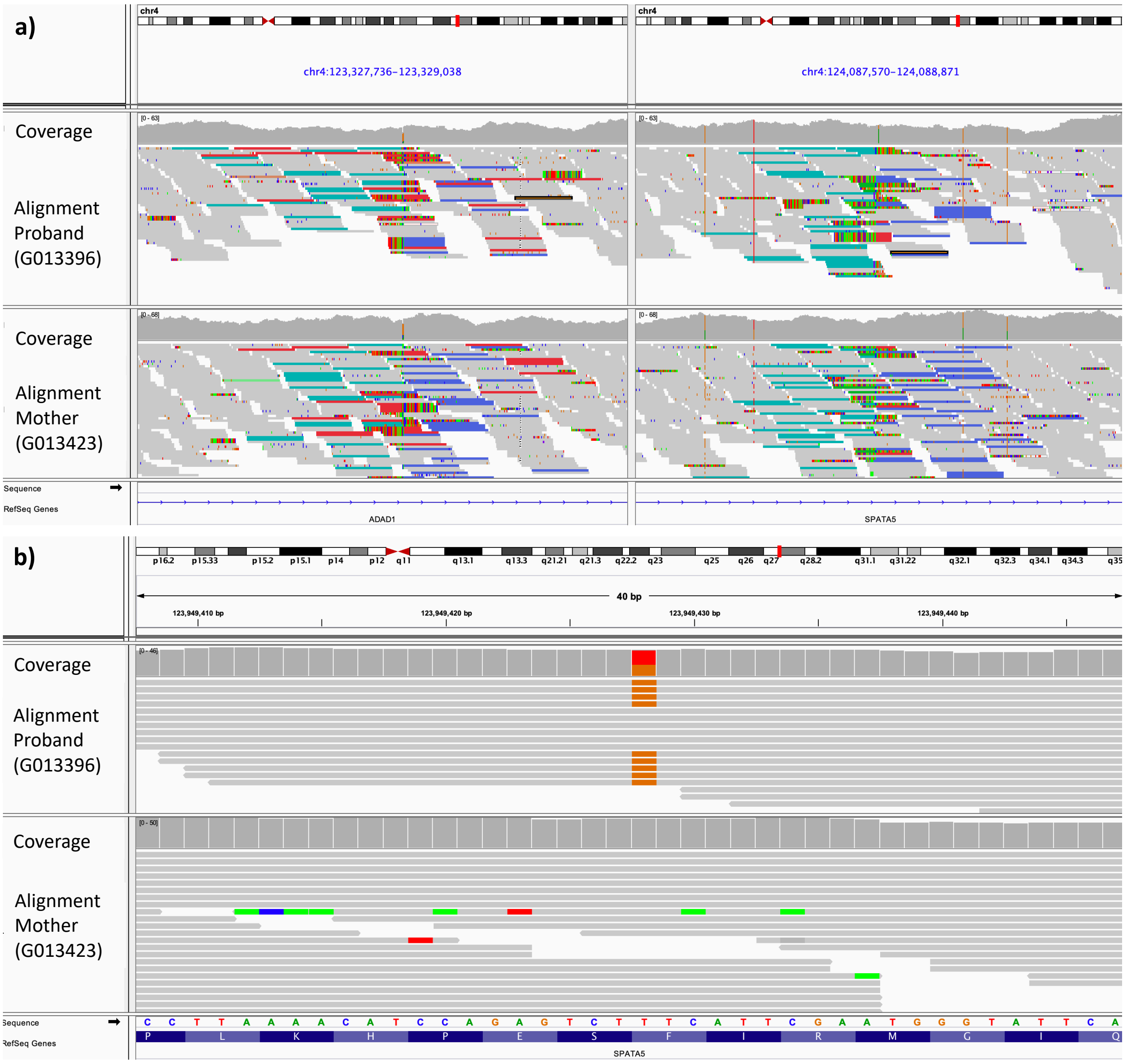

##### Case 2: G004131

Participant with Tuberous Sclerosis with no pathogenic/likely pathogenic (P/LP) variant identified in known Tuberous Sclerosis genes despite a long diagnostic odyssey. A heterozygous deep intronic variant in *TSC2* gene [MIM: 191092] was identified in this individual (Chr16:g.2127477G>A, ENST00000219476.3:c.2838-122G>A) in 17% (4/23) of the reads at this position, suggesting mosaicism (SFigure 3b). The variant is absent in gnomAD and has been reported before as associated with Tuberous Sclerosis (22). It has been previously shown that it creates a novel splice acceptor site leading to the insertion of 120 nucleotides upstream of exon 26, resulting in the premature truncation of the protein.

##### Case 3: G004703

Individual with ataxia, recurrent lactic acidosis and myopathy. This patient had the MT:3243A>G variant, which is a non-coding transcript exon variant in *MT-TL1* gene [MIM: 590050], in heteroplasmy at a level of 91% in blood. This is one of the most thoroughly studied and best characterized disease-causing mitochondrial (MT) variants, and is associated, amongst other phenotypes, with MELAS (myopathy, encephalopathy, lactic acidosis, and stroke like episodes) (23, 24), which was consistent with the patient's phenotype.

**SFigure 5.** MT variant identified in individual G004703 (MT:3243A>G) is present in 91% heteroplasmy.

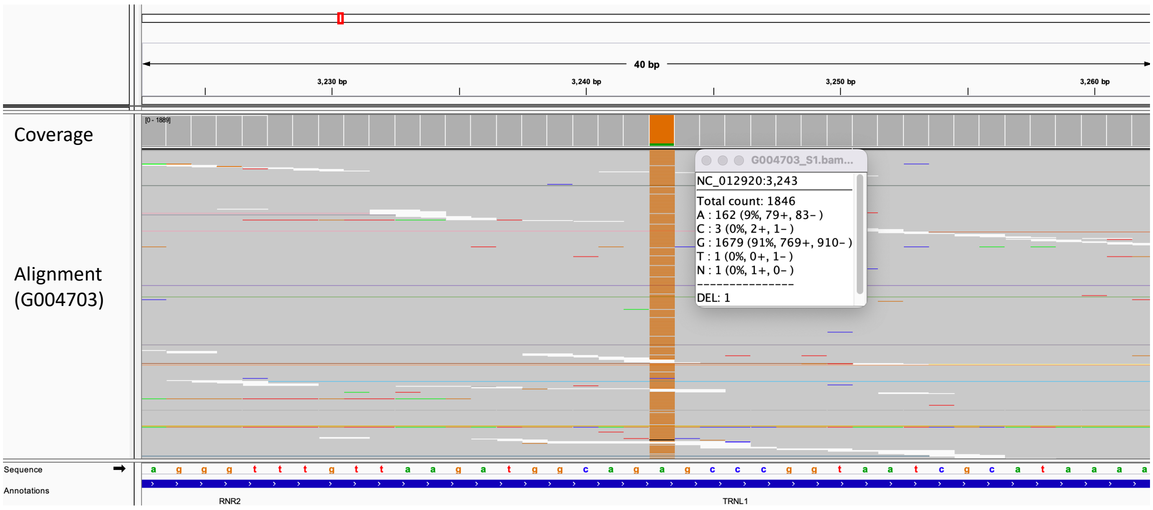

##### Case 4: G013808

Male individual with rod-cone dystrophy, ataxia and cerebellar atrophy. A heteroplasmic missense variant (MT:9032T>C, ENSP00000354632.2:p.Leu169Pro) in *MT-ATP6* gene [MIM: 516060] was observed at a level of 83% in blood. This variant has been previously associated with neurogenic muscle weakness, ataxia and retinitis pigmentosa (NARP) (25), consistent with the patient's phenotype.

**SFigure 6.** MT variant identified in individual G013808 (MT:9032T>C) is present in 83% heteroplasmy.

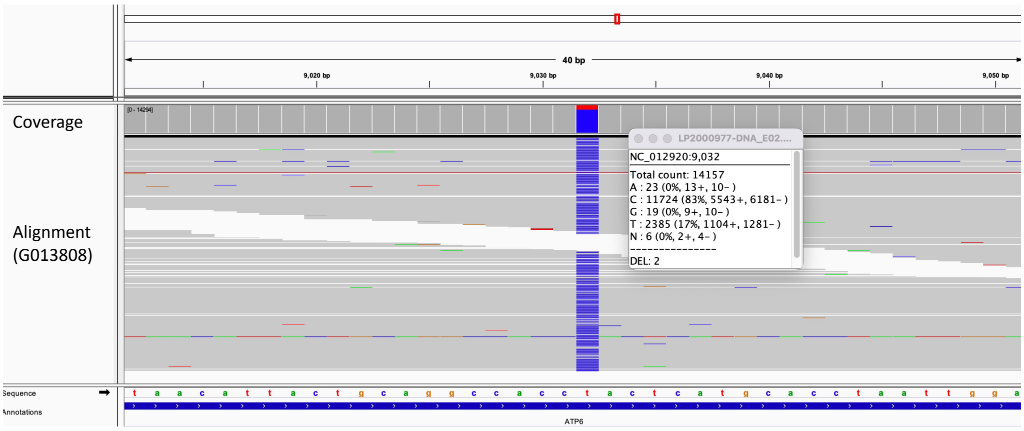


##### Case 5: G012198

Female individual with axial hypotonia and distal dystonia with a homoplasmy variant (ENSP00000354961.2:p.Arg340His) in *MT-ND4* gene [MIM: 516003]. This variant has been previously associated with Leber Hereditary Optic Neuropathy (26) and deemed to be likely pathogenic.

**SFigure 7.** MT variant identified in individual G012198 (MT:11778G>A) is present in homoplasmy.

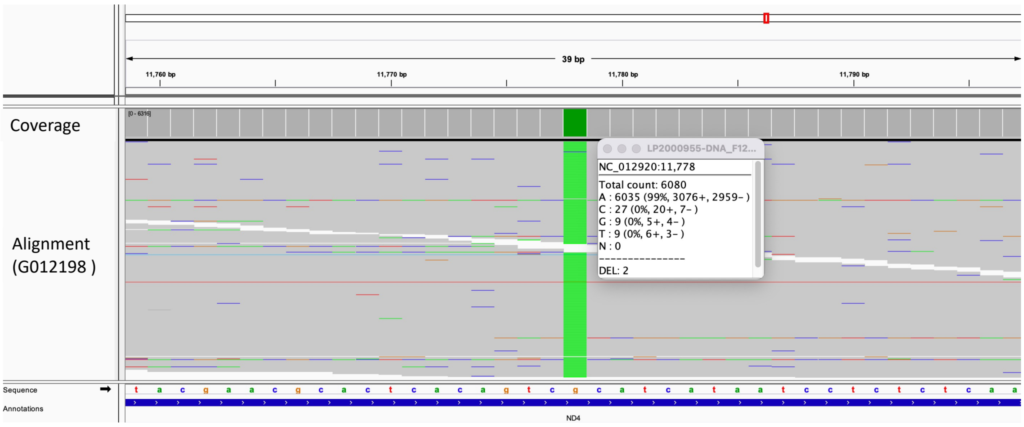


##### Case 6: NGC00375_01

Male with dystonia, learning difficulties and behavioural problems. This individual had a *de novo* complex SV disrupting *SGCE* [MIM: 604149], a gene associated with dystonia, which was deemed to be LP. Initially, multiple copy number losses in 7q21.3 were identified by microarray, including a loss at the 5' end of the gene *SGCE* but predicted to be intronic and not disrupting the gene.

However, short-read sequencing demonstrated the deletions included exons 1 and 11 of this gene (ENST00000648936.2) and showed that it was likely part of a complex SV. Resolution of the variant was attempted by short-read GS but it could not be achieved due to the presence of homology at the breakpoints. Long-read GS allowed SV characterization and resolved the complex rearrangement that involved 37 breakpoints between chromosomes 7, 10 and 12 (Figure 2A, STable 6).

##### Case 7: G012664

Male individual with paroxysmal dyskinesia and bulbar palsy. This case was sequenced as a trio and a complex rearrangement was suspected due to the presence of a high number of duplications across multiple chromosomes, inherited from the unaffected mother. Duplicated fragments involved the chromosome X and a X-linked mode of inheritance was suspected. However, the variant could not be resolved by short-read GS due to homology at the breakpoints. Long-read GS was used to determine the genomic architecture of this event, which resulted in a complex event involving 26 duplicated fragments of 24Kb median size (sd ± 12Kb) from 14 different chromosomes (Figure 2B). Although no protein coding gene was predicted to be disrupted, we couldn't rule out the possible regulatory effect of this event, and it was classified as VUS.

##### Case 8: G013407

Female with EIEE and a heterozygous missense variant in *DNM1* gene [MIM: 616346], which is associated with epileptic encephalopathy. The variant was not present in the unaffected father, but maternal DNA was not available. Given that 80% of *de novo* mutations occur in the paternal allele (27), we performed long-read GS to determine the haplotype of the variant and *de novo* status. Unfortunately, the closest informative SNV was 7,048 bp from this position and there were no reads of this length covering the region (average read length 6,723 +- 4,695). Therefore, the variant was classified as VUS.

**SFigure 8.** Heterozygous missense variant in *DNM1* in G013407 is absent in unaffected parent G013795.

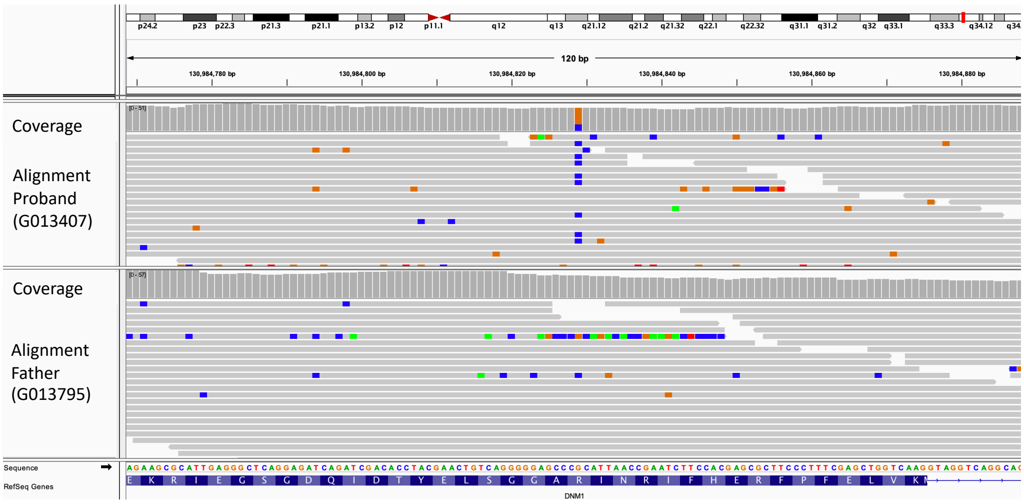

##### Case 9: G000973

Female with early onset dementia, spastic paraplegia and thin corpus callosum. Three deletions and two inversions were called by Manta algorithm in *KIF5C* [MIM: 604593], a gene associated with complex cortical dysplasia with other brain malformations, including thin corpus callosum, following autosomal dominant mode of inheritance. Manual inspection of the short-read GS alignment in IGV suggested the presence of a retroelement, with apparent deletions of introns 19-25 (ENST00000435030.1), with a similar structure than ENST00000482151.1, a transcript highly expressed in human brain (https://gtexportal.org/home/gene/ENSG00000168280). Discordant reads mapping to [GRCh37] Chr5:g.2,500,440 also supported the insertion of the retrotranscribed sequence in a L1MD sequence, possibly suggesting a LINE-L1 driven mechanism (Figure 2C). However, the two inversion calls ([GRCh37] Chr2:149,854,916-149,879,613 and Chr2:149,856,936-149,879,613) suggested a more complex event, and long-read GS was used to phase inversions with the retroelement. Long-reads facilitated phasing and showed that the inversion occurred at the 5' of the retroelement, which has previously seen in other retroelements (28), and *KIF5C* itself had not been disrupted. Although the insertion was not affecting any protein coding gene, it was classified as VUS since reports have shown that retroelements can interfere with gene expression by other mechanisms such as silencing by transcriptional or RNA interference (29).

##### Case 10: G013428

Individual with global developmental delay, hypotonia with movement disorder, sensorineural hearing impairment, microcephaly and delayed visual maturation with esotropia. An inversion involving *CASK* [MIM: 300172] was called in the short-read GS data (Manta call [GRCh37] ChrX:41,426,631-41,501,873). However, quality of the alignment at the breakpoints was poor due to the presence of LINE retroelements and sequence homology (Figure 2D), and the variant couldn't be confirmed by long-range PCR due to low sequence complexity. We therefore sought to validate it using long-read GS, and no inversion was called involving *CASK* gene. Manual inspection of the long-read alignment in IGV also supported the absence of the event and the variant was classified as unlikely to be real.

**SFigure 9. Long-read GS sequencing quality control** results from five individuals. **A)** Median coverage of 14.6x was observed across individuals. **B)** Read length. **C)** Proportion of the genome covered at specific minimum coverage. **D)** Number of SVs by median coverage. **E)** Number of SVs by sample and type. **F)** Number of SVs by size.

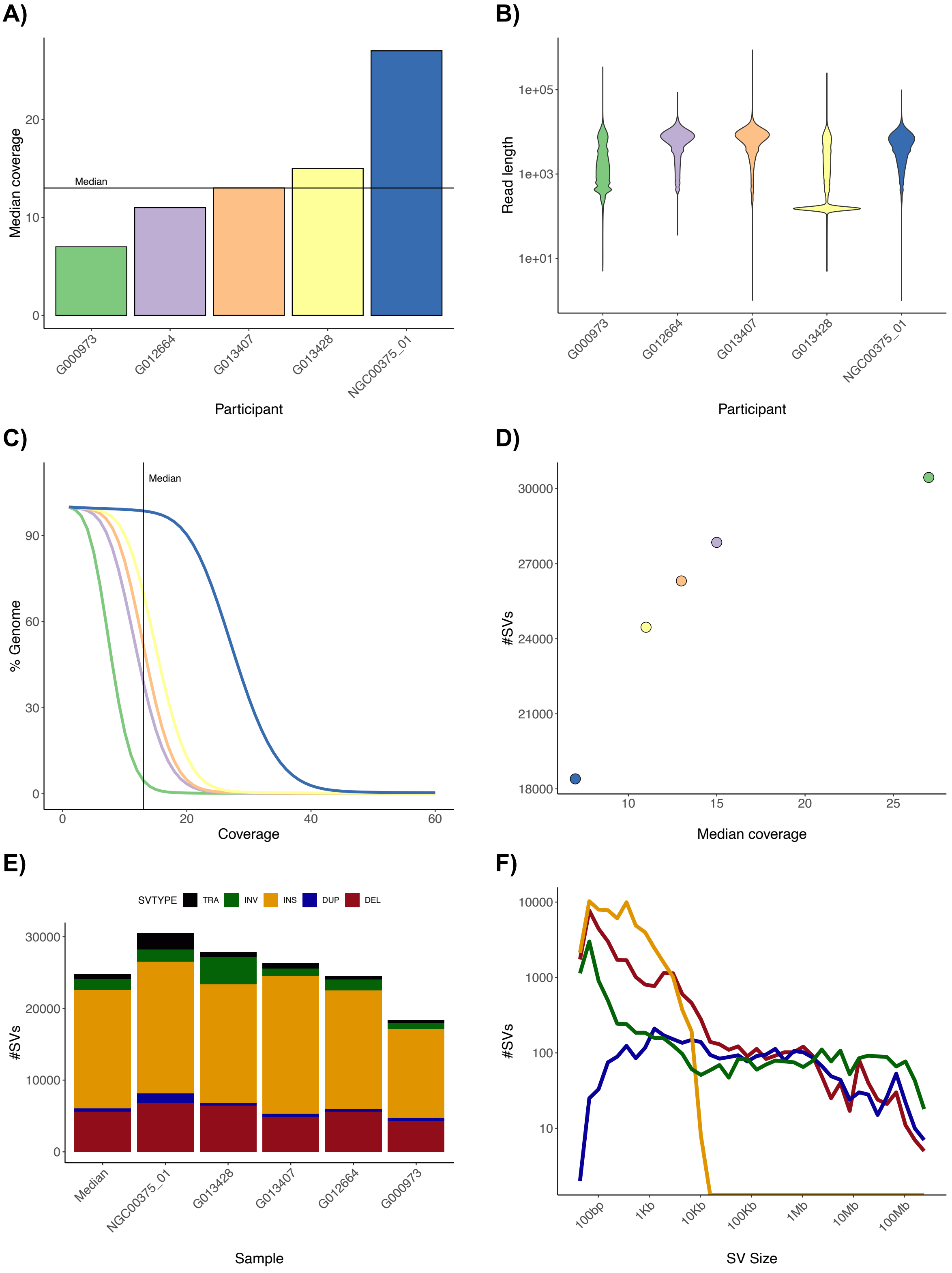
